# *Neisseria gonorrhoeae* MtrCDE Efflux Pump During *In Vivo* Experimental Genital Tract Infection in Men and Mice Reveals the Presence of Within-Host Colonization Bottleneck

**DOI:** 10.1101/2023.06.23.23291824

**Authors:** Andreea Waltmann, Jacqueline T. Balthazar, Afrin A. Begum, Nancy Hua, Ann E. Jerse, William M. Shafer, UNC-Global Clinical Trials Unit/DMID 09-0106 Study Team, Marcia M. Hobbs, Joseph A. Duncan

## Abstract

The MtrCDE efflux pump of *Neisseria gonorrhoeae* exports a wide range of antimicrobial compounds that the gonococcus encounters at mucosal surfaces during colonization and infection. Here, we evaluate the role of this efflux pump system in strain FA1090 in human male urethral infection with a Controlled Human Infection Model. Using the strategy of competitive multi-strain infection with wild-type FA1090 and an isogenic mutant strain that does not contain a functional MtrCDE pump, we found that the presence of the efflux pump during human experimental infection did not confer a competitive advantage. This finding is in contrast to previous findings in female mice, which demonstrated that gonococci of strain FA19 lacking a functional MtrCDE pump had a significantly reduced fitness compared to the wild type strain in the lower genital tract of female mice. We conducted competitive infections in female mice with FA19 and FA1090 strains, including mutants that do not assemble a functional Mtr efflux pump, demonstrating the fitness advantage provided byt the MtrCDE efflux pump during infection of mice is strain dependent. Our data indicate that new gonorrhea treatment strategies targeting the MtrCDE efflux pump functions may not be universally efficacious in naturally occurring infections. Owing to the equal fitness of FA1090 strains in men, our experiments unexpectedly demonstrated the likely presence of an early colonization bottleneck of *N. gonorrhoeae* in the human male urethra.

**TRIAL REGISTRATION:** Clinicaltrials.gov NCT03840811.

## Introduction

*Neisseria gonorrhoeae* is exquisitely adapted to humans, its only natural host. The gonococcus is equipped with multiple mechanisms that allow it to quickly respond to host innate immune responses. The multiple transferrable resistance (Mtr) phenotype was first identified in 1973 as providing decreased susceptibility of gonococci to structurally diverse hydrophobic antibiotics, detergents and dyes (1) and was postulated to be due to decreased permeability of the bacterial outer membrane to these antimicrobials (2). Over two decades later it was shown that the resistance phenotype was due to the relief of transcriptional repression of the *mtrCDE* –encoded efflux pump operon. The MtrCDE efflux pump is a member of the resistance-nodulation-division family possessed by many Gram-negative bacteria (3, 4). The MtrCDE pump is critical to the ability of *N. gonorrhoeae* to export a wide range of substrates that bathe mucosal surfaces, including host-derived antimicrobials (antimicrobial peptides, bile salts, progesterone, fatty acids, and cathelicidins, such as human LL-37 or its murine homologue CRAMP-38), and antibiotics (3, 5–10). The Mtr system has also been associated with enhanced resistance to killing by human neutrophils and neutrophil products (11).

The *mtrCDE* operon is transcriptionally regulated by *cis*- and *trans*-acting control processes (5, 12, 13). MtrD is the inner membrane component of the pump and is connected to MtrE in the outer membrane through the periplasmic membrane fusion lipoprotein MtrC (4, 6, 10). Efflux is dependent on energy supplied by the proton motive force transduced by MtrD (14, 15). Expression of *mtrCDE* in wild type gonococci is subject to repression by MtrR (7, 16, 17) and, in the presence of an inducer, activation by MtrA (17, 18). Loss-of-function *mtrD* mutations significantly reduce the ability of gonococci to colonize the lower genital tract of female mice in experimental infection (19). Higher levels of the pump, on the other hand, either through induction by MtrA, or de-repression through either loss of MtrR or through mutations in the Mtr DNA-binding region enhance resistance to antibiotics (8, 20–23), host-derived antimicrobials (8), and *in vivo* fitness in the mouse infection model (24). In strains that overexpress the pump, loss of MtrCDE can increase antibiotic susceptibility in otherwise clinically-resistant gonococci (25), including strain H041, which was isolated from the first reported extended-spectrum-cephalosporin resistant case of gonorrhea (26). These findings support the idea that the MtrCDE efflux pump is important during *in vivo* infection (8, 24).

We hypothesized that mutations that abrogate the expression of the Mtr efflux pump would affect *N. gonorrhoeae* infectivity in experimentally infected human males. We and others have developed a model of experimental human gonococcal infection (reviewed in (27, 28)), which takes advantage of the self-limited nature of gonococcal urethral infection in men. Infection can be initiated in male participants through intraurethral inoculation, and this model has proven to be a safe and efficient means of studying human urethral gonococcal infection without serious complications (27, 28). With this model and the strategy of competitive infections initiated by a mixture of wild type strain FA1090 and an isogenic FA1090 mutant that lacks MtrD, we sought to measure the importance of the MtrCDE efflux pump on N. gonorrhoeae during experimental urethral infection of human males.

## Results

### Infectivity and clinical courses of infection in non-competitive infections

The genotypes of inoculating strains are shown in Table 1. Results are summarized in Table 2. The evaluable population included participants who received a dose of *N. gonorrhoeae* within 1 log of the intended dose of one million organisms [i.e. Log_10_(5), below which the inoculum size is considered low at approximately ID_50_] and reached an objective study endpoint (urethral discharge or day 5). Six male participants were inoculated with wild type FA1090 A26 alone and included in downstream analyses. Of nine men inoculated with FA109011*mtrD* alone, two were excluded: one volunteer received a low inoculum dose at approximately ID_50_ [inoculum size=Log_10_(5.0)] and a second volunteer requested treatment without signs, symptoms, or positive cultures on day 2 post inoculation. Among participants included in final analyses, there were no significant differences between the two strains with respect to infectivity, inoculum size, pyuria on treatment day, and time to treatment (Table 2). Among evaluable and infected volunteers, bacteriuria on treatment day did not differ significantly between the two strains (Table 2), nor were they deemed to be clinically meaningful. These results indicate the MtrCDE efflux pump is not required for urethral infection in experimentally infected men.

**Table 1.** Bacterial strains used in this study.

| Strain | MtrD pump expression | Reference |
| --- | --- | --- |
| FA1090 A26 | Not inducible due to 11 bp deletion in coding region of activator *mtrA* | (39) |
| FA1090Δ*mtrD* | None | This study |
| FA19 | Inducible (wild type *mtrA*) | (86) |
| FA19Δ*mtrD* | None | This study |
| FA19 *mtrD::kan* | None | This study and (19) |

**Table 2.** Clinical course parameters of non-competitive infections in men inoculated with *N. gonorrhoeae* FA1090 or *N. gonorrhoeae* FA1090*11mtrD*.

| Participant | Strain | Inoculum size<sup>a</sup> | Infected? | Bacteriuria<sup>b</sup> | Pyuria<sup>c</sup> | Day of Treatment<sup>d</sup> |
| --- | --- | --- | --- | --- | --- | --- |
| 232 | FA1090 | 6.6 | Y | 6.4 | 6.7 | 3 |
| 241 |  | 6.8 | Y | 4.5 | 6.3 | 2 |
| 242 |  | 6.7 | Y | 6.3 | 7.0 | 3 |
| 243 |  | 6.8 | Y | 5.5 | 7.1 | 3 |
| 251 |  | 5.9 | Y | 2.0 | 6.4 | 1 |
| 252 |  | 6 | Y | 6.0 | 6.9 | 4 |
| Summary<sup>e</sup> |  | 6.5 (6.0-6.9) | 100% (54%, 100%) | 4.8 (3.0-7.6) | 6.7 (6.4-7.1) | 3.0 (2-3) |
| 216 | FA1090Δ *mtrD* | 5.8 | Y | 6.2 | 6.2 | 3 |
| 220 |  | 6.4 | Y | 3.2 | 6.8 | 1 |
| 222 |  | 6.7 | Y | 4.0 | 8.0 | 2 |
| 223 |  | 6.7 | Y | 6.0 | 8.0 | 3 |
| 271 |  | 6.0 | Y | 0.7 | 6.0 | 4 |
| 272 |  | 6.1 | N | ND | 6.0 | 5 |
| 273 |  | 6.1 | Y | 2.0 | 6.9 | 4 |
| Summary<sup>e</sup> |  | 6.2 (5.9-6.6) | 86% (42%, 100%) | 3.0 (1.2-7.0) | 6.8 (6.1-7.6) | 3.0 (2-4) |
| *p-value*<sup>g</sup> |  | 0.39 | 0.54 | 0.23 | 0.77 | 0.50 |

Y=yes, N=no
<sup>a</sup>Log<sub>10</sub> CFU *N. gonorrhoeae* mixture delivered to intraurethrally
<sup>b</sup>Log<sub>10</sub> CFU *N. gonorrhoeae* recovered/mL urine sediment on day of treatment
<sup>c</sup>Log<sub>10</sub> wbc/mL urine sediment on day of treatment
<sup>d</sup>Days from inoculation to antibiotic <sup>e</sup>Summary statistics refer to geometric means with 95% confidence intervals shown in brackets, except for the infectivity summary, which denotes the percentage of participants deemed to have been infected (with 95% confidence intervals), and time to treatment, which denotes the median time from inoculation to treatment (with interquartile range). P-values were generated using the two-sided two-sample Wilcoxon rank-sum test, except for the infectivity comparison for which the one-sided Fisher's exact test was used.

### The MtrCDE efflux pump does not confer a fitness advantage during urethral infection of men experimentally infected with strain FA1090 of *N. gonorrhoeae*

To determine whether the FA1090 Mtr efflux pump confers a fitness advantage to *N. gonorrhoeae* in the male urethra, we conducted competitive infections using mixed FA1090 and FA1090Δ*mtrD* inocula. Twelve participants in four cohorts were inoculated with strain mixtures. The evaluable population for competitive index calculations included participants who received a dose of *N. gonorrhoeae* within 1 Log_10_ of the intended dose (Log_10_ 6) and reached an objective study endpoint (urethral discharge or day 5) for whom a competitive index (CI) could be calculated. Ten participants were evaluable and included in competitive fitness evaluations (Table 3). One volunteer was urine culture-negative on treatment day, and thus the CI could not be calculated. A second volunteer did not comply with specimen collection requirements, because of which the processed urines were not the first void of the day. Among evaluable participants, infectivity and course of infection parameters were similar to those in non-competitive infections (Table 3).

**Table 3.** Infectivity and course of infection parameters in men infected competitively with FA1090+FA1090Δ*mtrD,* and comparisons to the same parameters from non-competitive infections in men.

| Participant | Strain | Inoculum size<sup>a</sup> | Infected? | Bacteriuria<sup>b</sup> | Pyuria<sup>c</sup> | Day of Treatment<sup>d</sup> |
| --- | --- | --- | --- | --- | --- | --- |
| 281 | FA1090+<br>FA1090Δ<br>*mtrD* | 6.6 | Y | 2.9 | 7.1 | 2 |
| 291 |  | 5.8 | Y | 5.2 | 6.9 | 4 |
| 292 |  | 5.8 | Y | 5.0 | 6.5 | 4 |
| 300 |  | 6.5 | Y | 4.6 | 6.9 | 4 |
| 301 |  | 6.5 | Y | 5.1 | 6.5 | 3 |
| 302 |  | 6.6 | Y | 5.0 | 6.3 | 2 |
| 303 |  | 6.5 | Y | 5.0 | 6.6 | 4 |
| 311 |  | 6.5 | Y | 3.2 | 6.9 | 2 |
| 313 |  | 6.3 | Y | 3.5 | 6.8 | 4 |
| 314 |  | 6.4 | Y | 3.5 | 7.0 | 4 |
| **Summary<sup>e</sup>** |  | **6.3 (6.1-6.6)** | **100% (69%, 100%)** | **4.2 (3.6-4.9)** | **6.7 (6.6-6.9)** | **4 (2.0-4.0)** |
| **Summary** | FA1090 | **6.5 (6.0-6.9)** | **100% (54%, 100%)** | **4.8 (3.0-7.6)** | **6.7 (6.4-7.1)** | **3.0 (2.0-3.0)** |
| **Summary** | FA1090Δ<br>*mtrD* | **6.2 (5.9-6.6)** | **86% (42%, 100%)** | **3.0 (1.2-7.0)** | **6.8 (6.1-7.6)** | **3.0 (2.0-4.0)** |
| ***p-value<sup>f</sup>*** |  | **0.38** | **0.56** | **0.25** | **0.91** | **0.48** |

Y=yes, N=no
<sup>a</sup>Log<sub>10</sub> CFU *N. gonorrhoeae* mixture delivered intraurethrally
<sup>b</sup>Log<sub>10</sub> CFU *N. gonorrhoeae* recovered/mL urine sediment on day of treatment
<sup>c</sup>Log<sub>10</sub> wbc/mL urine sediment on day of treatment
<sup>d</sup>Days from inoculation to antibiotic treatment
<sup>e</sup>Summary statistics refer to geometric means with 95% confidence intervals shown in brackets, except for the infectivity summary, which denotes the percentage of participants deemed to have been infected (with 95% confidence intervals) and time to treatment, which denotes median time from inoculation to treatment (with interquartile range). P-values were generated using Kruskal-Wallis tests, except for the infectivity comparison for which the Fisher's exact test was used.

The proportion of each strain in the inoculum and in urine sediment collected from each participant on treatment day was determined by sub-culturing up to 96 colonies from each positive urine sediment culture and enumerating strain-specific colony-forming units (cfu)by real-time colony PCR (Supplementary Table 2). To assess whether the fitness of the FA1090Δ*mtrD* mutant was different than that of wild type, the ratio of mutant cfu as identified by colony real-time PCR was compared to wild-type cfu for up to 96 recovered colonies per participant. Log_10_(CI) of participants inoculated with a mixture of FA1090 and FA1090Δ*mtrD* group were compared using a Wilcoxon one-sample signed rank test to a hypothetical median of 0. We observed equal outcomes favoring the mutant strain and the wild type strain and overall there was not enoughevidence to reject the null hypothesis (*p=* 0.477, one-sample signed rank test, Figure 1A). Thus, expression of the FA1090 Mtr efflux pump did not confer a competitive advantage during experimental human urethral infection.

**Figure 1.**
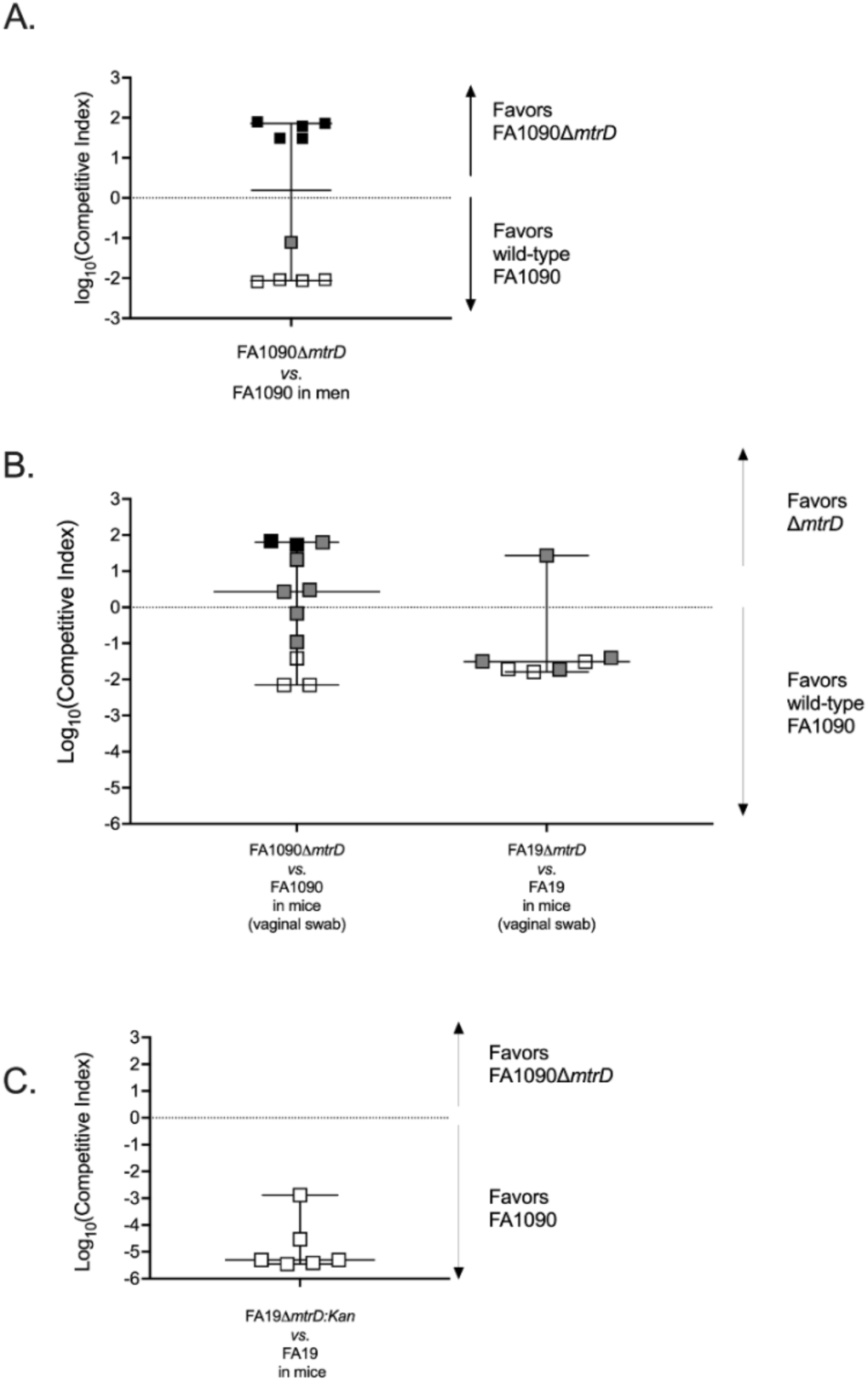
Results of competitive infections in men (n=10) and female mice. **A.** Ten men were competitively infected with mixtures of FA1090 and FA1090Δ*mtrD.* Competitive indices were calculated as cfu mut OUT/cfu wt OUT ÷ cfu mut IN/cfu wt IN recovered from first-void urine of experimentally infected men. CIs are graphed on the logarithmic scale. Thus, Log_10_CI greater than 0 indicated the mutant was favored and Log_10_CI less than 0 indicates that the wild type was favored. For men, the last day of positive cultures was treatment day when clinical urethritis was detected and/or when antibiotic treatment was given. Horizontal bars are the median CI for each group. White squares refer to men from whom only the wild type was recovered the day of treatment. Grey squares denote men from whom a mix of the two strains was recovered. Black squares denote men from whom only the mutant strain was recovered. **B.** Eleven mice were inoculated with mixtures of FA1090 and FA1090Δ*mtrD* and 7 mice with mixtures of FA19 and FA19Δ*mtrD*. CIs were calculated using the same formula as for the men using the cfu counts from mouse vaginal swabs on the last day of positive cultures. CIs are graphed on the logarithmic scale. Horizontal bars are the median CI for each group. White squares refer to mice from whom only the wild type was recovered on last day of positive cultures. Grey squares denote mice from whom a mix of the two strains was recovered. Black squares denote mice from whom only the mutant strain was recovered. **C.** Six mice were inoculated with mixtures of FA19 and FA19*mtrD::kan* and CIs calculated using cfu counts from mouse vaginal swabs on the last day of positive cultures. CIs are graphed on the logarithmic scale. Note the different y axis scale for panel C, imposed by the lower limit of detection of quantitative selective culture on GC agar for the FA19+FA19 mtrD::kan competitive infections.

### The *in vivo* fitness advantage of the Mtr efflux pump is strain dependent in female mice

Given that previous studies on FA19 *mtr* mutant strains in female mice showed that *mtrD* and *mtrE* mutants were attenuated for murine vaginal infection, and that the Mtr pump is important for *in vivo* survival, either through induction by MtrA, the activator of the pump expression, or de-repression through loss of MtrR, the repressor of pump expression (8, 19, 24), the results of the human competitive infections with FA1090 and FA1090Δ*mtrD* were unexpected. Furthermore, when investigating the role of LptA in FA1090 strains, which also increases gonococcal resistance to innate defenses such as cationic antimicrobial peptides, the two models showed similar results (29). Experimental infection of female BALB/c mice has been used extensively to study *N. gonorrhoeae* pathogenesis (8, 19, 24, 30–37). To determine whether this discrepancy reflected differences between model systems, between strains, or between mutant generation methodologies, we conducted competitive infections in female mice with the following strain mixtures of wild type and mutant: wild type FA1090 with FA1090Δ*mtrD,* wild type FA19 with FA19Δ*mtrD*, and wild type FA19 with FA19*mtrD::kan*, which was tested previously (19). For this study, groups of female BALB/c mice were vaginally inoculated with mixtures containing similar numbers of wild type and mutant bacteria (total dose, Log_10_ 5.8-6). The number of colonies of each strain recovered from vaginal swabs was determined for up to 5 days after inoculation; strain determination for each cfu was done by real-time PCR for competitive infections with FA1090 and FA1090Δ*mtrD* and with FA19 and FA19Δ*mtrD*; quantitative selective culture was used for FA19 and FA19*mtrD::kan* competitive infections.

Among the 11 mice competitively infected with FA1090 and FA1090Δ*mtrD* (Supplementary Table 3), we observed five outcomes that favored the wild type FA1090 and six outcomes that favored FA1090Δ*mtrD* (*p=*0.672 1-sided Fisher’s exact, Figure 1B). There was no significant departure from the null hypothesis as the Log10(CIs) were not from different from the hypothetical median of 0 (*p=*0.943, Wilcoxon one-sample signed rank test). Results with FA1090 strains were not different in the two model systems (*p=*0.441, Mann-Whitney rank sum test), although more mice than men had mixed strain recovery on the final positive culture day (mice: 6 /11, 54.5%; men: 1/10, 10.0%, *p=*0.045 one-sided Fisher’s exact).

Among the 7 mice infected with mixtures of FA19 and FA19Δ*mtrD,* all but one outcome favored wild type FA19, with a significant difference from the hypothetical median of 0 (*p=*0.031, Wilcoxon one-sample signed rank test, Figure 1B). The FA1090 and FA19 results were significantly different in the mouse model (*p=*0.047, Mann-Whitney rank sum test). All outcomes in the 6 mice competitively infected with FA19 and FA19*mtrD:*:kan favored the wild type, with no mixed strain recovery on final day of positive cultures (*p*=0.030, Wilcoxon one-sample signed rank test, Figure 1C) in line with previously published results (8, 19, 24).

We conclude that the *in vivo* phenotype of the FA1090 mutant that does not produce the MtrCDE pump is the same in the male urethritis and female mouse models, but that an interesting strain difference between FA1090 and FA19 mutants was detected in the mouse model.

### Strain dynamics during human infection indicate the presence of a colonization bottleneck

Lack of a functional Mtr efflux pump did not alter the fitness of the isogenic mutant relative to wild type FA1090. Thus, our strategy of competitive infection enabled the incidental observation of a potential urethral colonization bottleneck. Competitive infections with strains of equal fitness have been used to study bottlenecks in other infection models. A bottleneck refers to barriers to colonization or infection that substantially reduce the original population size in an inoculum, such that a lower number of surviving organisms establish colonization and replicate in the host. Our findings indicate that such a bottleneck exists early in the course of gonococcal infection.

Figure 2A depicts the theoretical outcomes of one individual infection initiated with a 50:50 mixture of two hypothetical strains under conditions of bottleneck and absent bottleneck. When a bottleneck is not present, the strategy of competitive infections can have two outcomes: if both strains have equal fitness, the bacteria recovered from the host after inoculation mirrors the inoculum (outcome 1). If one of the strains has an advantage, the expectation is for that strain to progressively take over the dominate over the gonococcal population over course of infection (outcome 2). Under the scenario of a bottleneck, if the same strain is at a competitive advantage, then it is expected to dominate from early until late infection (outcome 3). When neither has a fitness advantage and a bottleneck is preset, both strains will have equal chance of passing through the bottleneck and establishing an infection; therefore, in outcome 4, equal numbers of competitive infections would be dominated by each strain.

**Figure 2.**
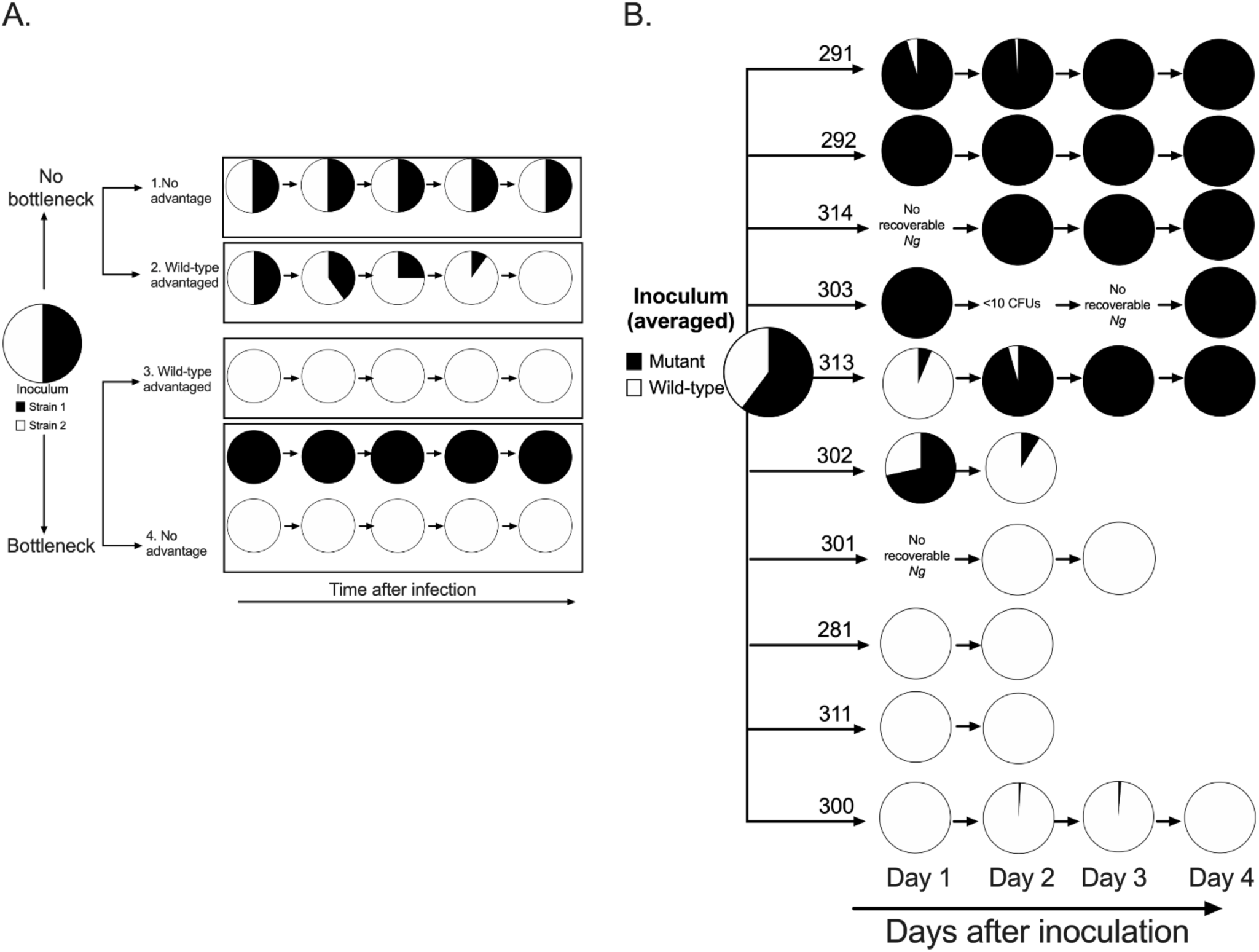
Gonococcal population dynamics during the early course of infection. A) Theoretical infection outcomes with respect to strain composition for infections initiated from 50:50 mixtures of two strains, shown in black and white, that differ in their competitive advantage in scenarios of bottleneck present early in the infection or no bottleneck (adapted with permission from (27). Each pie chart represents a gonococcal population containing predicted proportions of the two theoretical strains. When the two strains have equal fitness and a bottleneck is present early in the course of infection that results in a population size restriction, we predict that equal numbers of outcomes favoring each strain will be observed. B) Empirically observed outcomes in the current study for infections initiated with mixed inocula. Two to four men were inoculated with the same inoculum preparation in each of the four study cohorts. A total of 10 evaluable participants were available for final analysis (281, 291, 292, 300, 301, 302, 303, 311, 313, 314). The pie chart representing the average inoculum from the four cohorts refers to the mean strain composition of the four independent inocula and on average comprised of 60% mutant strain and 40% wild type. Each participant pie chart represents the daily mixture of gonococcal strains recovered from the urine over the 1- to 5-day period after inoculation. Last pie chart in the series for each participant refers to the study treatment day when clinical urethritis was apparent and/or when antibiotic treatment was administered.

Figure 2B describes the observed outcomes for urine culture positive days in each of the 10 evaluable human participants, who on average received 40:60 mixed inocula of FA1090 and FA1090Δ*mtrD*. In the majority of men (8/10, 80.0%, Figure 2B), the predominant FA1090 strain on treatment day began dominating early in the infection (day 1 post-inoculation). In 2/10 men (20.0%), the predominating FA1090 strain from treatment day was different than the strain dominating the recovered *N. gonorrhoeae* cfu from day 1 post-inoculation. In all but one man (9/10, 90.0%), the strain composition of the first void urine on the final study day was uniformly dominated by either strain. Strain dynamics from urine culture positive days support the gonococcal bottleneck hypothesis.

## Discussion

In the female mouse model of gonorrhea, the MtrCDE efflux pump was found to be important to *in vivo* infection with *N. gonorrhoeae* strain FA19 (8, 19, 24). We anticipated human competitive infections with wild type FA1090 and an isogenic polar mutant of FA1090 that lacked a functional MtrCDE efflux pump to yield similar results. Contrary to our hypothesis, the two strains exhibited equal fitness. The previous investigations using the murine model were conducted with wild type strain FA19 and FA19 mutants whose MtrCDE efflux pump was rendered non-functional via insertional inactivation (8, 19, 24). Thus, the discrepancy between human and mouse results could be a reflection of strain, model, or mutagenesis methodology differences. To separate potential differences with respect to strain, we conducted competitive infections in mice with both FA1090 and FA19 parent strains and corresponding mutants that lacked a functional MtrCDE efflux pump due to clean deletions of the *mtrD* gene (FA1090Δ*mtrD* and FA19Δ*mtrD)*. The FA1090 strains used in mouse infections were the same strains used in human infections. The outcomes of competitive infection with FA1090 strains did not differ between two model systems, indicating that the Mtr pump is not required for colonization of murine lower genital tracts or human male urethras. However, mouse competitive infections with FA19 strains showed that FA19 wild type had significant competitive advantage relative to FA19Δ*mtrD*. This competitive advantage was maintained when tested against the same insertionally inactivated *mtrD* mutant used in the previous report (19) Nevertheless, differences between infection models and mutagenesis strategies were noted, namely the more frequent recovery of both wild type and mutant strains from mice compared to men during competitive infections with FA10190 strains, and from mice inoculated with wild type FA19 in combination with FA19Δ*mtrD* relative to mice inoculated with wild type FA19 and FA19*mtrD*::kan. Overall, our findings indicate that the fitness advantage conferred by the MtrCDE efflux pump in the murine infection model is strain-dependent. Two established lines of evidence suggest that strain dependent differences in MtrCDE-mediated enhanced fitness may also be present during human infection. First, FA1090 naturally expresses reduced levels of the pump compared to other strains, including FA19, due to an 11 bp deletion in the coding region of *mtrA* that results in loss of MtrA-mediated gene activation due to premature truncation of this transcriptional activator (18, 23). Thus, the null mutation in the *mtrD* gene in FA1090 may not result in a strong fitness phenotype compared to the parental FA1090 strain that already expresses a low level of MtrCDE (*i.e.,* low expression in the wild type *vs*. no expression in the mutant). This is consistent with the observed equal fitness of the strains during human and mouse competitive infections. Second, a recent compelling study that investigated the population genomics of the *mtrCDE* operon and its regulatory genes *mtrA* and *mtrR* using almost 5,000 global *N. gonorrhoeae* genomes, found that loss-of-function mutations related to the expression of the MtrCDE efflux pump are relatively common in nature (38). In this global meta-analysis (38), 362/4842 isolates (7.48%) were found to not have an inducible MtrCDE efflux pump due to mutations in *mtrA*, of which the vast majority (357/362, 98.6%) harbored the same 11-bp deletion as FA1090 *mtrA*. FA1090 was originally isolated from the endocervix of a female with probable disseminated gonococcal infection (39) and the global population genomics study found that loss of function mutations were over-represented among genomes from cervical isolates; 25 of 129 cervical isolates (19.4%) carried *mtrA* loss of function mutations compared to 61 out of 2,248 urethral isolates (2.71%) (38). This finding indicates that strains with low levels of MtrCDE efflux pump expression colonize the genital tracts of both females and males. Thus, it may be reasonable to conclude that the possibility of strain-dependent requirement of the MtrCDE efflux pump is representative of other naturally occurring gonococcal strains with non-inducible MtrCDE efflux pumps. Although the pump may not be required to establish infection in humans, evolving evidence suggests that overproducers of the pump may be better positioned for antimicrobial resistance, as position 823 in MtrD is predicted to be a site for binding antimicrobials (38, 40).

The equal fitness of FA1090 and FA1090Δ*mtrD* in urethral infection observed using competitive infection enabled the identification of a within-host bottleneck during the gonococcal infection process. To our knowledge, this is the first demonstration that productive infection with *N. gonorrhoeae* is likely initiated by a restricted number of gonococci rather than through a uniform expansion of the initial mixed inoculum, though the concept of a gonococcal colonization bottleneck was previously proposed (27). Transmission or colonization bottlenecks are drastic reductions in population size followed by clonal expansion of the “founder” organisms that initiate the productive infection. Bottlenecks have been observed in enteric and systemic infections caused by pathogenic bacteria in humans (41–47). Among sexually-transmitted infections, human immunodeficiency virus 1 (HIV-1) bottlenecks have been studied but to date no studies have demonstrated their presence in bacterial sexually transmitted infections (48–51).

We tracked the daily strain dynamics during human competitive infections from day 1 post-inoculation until the day of clinical urethritis (Figure 2). We noted that the “winning” strain outcompeted the “losing” strain within the first 24h of inoculation in the majority of participants (8/10) and persisted as dominant strain until the final study day. However, both strains were recovered from 8/29 positive cultures of 4/10 volunteers, albeit at very different proportions. Interestingly, in 2 participants (302 and 313) in which both strains were recovered, the strain dominance observed on the first day after inoculation reversed the following day. While the study was not designed *a priori* to study populaton bottlenecks during *in vivo N. gonorrhoeae* infection, our findings allude to some possible bottleneck characteristics with respect to type (selective or non-selective), size (how many organisms are able to breach the initial barrier?), and timing.

First, it is possible that the gonococcal bottleneck is non-selective and strains overcome the bottleneck with equal probability, as observed in murine models of *Streptococcus pneumoniae* infection (43, 53–56). Our imposed genetic diversity of two strains is insufficient to answer this question with any certainty. A selective bottleneck has also been described using *in vitro* selection with human serum and in baby rat models of infection with the only other pathogenic neisserial species, *Neisseria meningitidis* which happens to share 80-90% of its genome with the gonococcus (61). The study showed experimentally that even when controlling for inoculum size a strongly selective bottleneck for survival of meningococci is generated by bactericidal antibody directed against the carbohydrate antigen produced by the phase variable enzyme LOS glycosyltransferase G*, lgtG* (61). Meningococcal variants existing within a population that do not express *lgtG* survive when exposed to a bactericidal monoclonal antibody specific for the epitope, whereas isogenic “ON” variants are killed essentially selecting for minority *ltgG* “OFF” variants. Similar dynamics were observed in an infant rat model given the monoclonal antibody, with *ltgG* “OFF” variants escaping into the bloodstream after intraperitoneal inoculation. Interestingly, the same gene also exists in *N. gonorrhoeae* (LOS glycosyltransferase G*, lgtG*) (62) and it too undergoes phase variation (63) and is the target of a monoclonal antibody with promising immunotherapeutic potential (64, 65).

While the exact gonococcal bottleneck size cannot be inferred from our data, the recovery of both strains from the same urine specimen indicates that the bottleneck is wider than one single cell. This would stand in contrast to bottlenecks for other human bacterial pathogens, which have tight single-cell bottlenecks (43, 45, 66) or for HIV-1 in which 80% of HIV-1 heterosexual transmission is initiated from a single HIV-1 genetic variant (48–51), reviewed in (47).

Finally, based on the data in the current study, the timing of the bottleneck may vary between individuals and/or there may be more than one sequential bottleneck in play over the first 48 hours after exposure to *N. gonorrhoeae*. The timing of urine sampling in our experiments (every 24h) is likely too infrequent to precisely determine the timing of the bottleneck. However, previous studies on the kinetics of opacity protein selection and pilin antigenic variation during experimental human infection with FA1090 support the idea that the infecting population encounters strong selective forces early in the infection process (67–70). Opacity and pilin proteins are known for their extensive phase and antigenic variation in *N. gonorrhoeae*. The opacity and pilin protein repertoire relative to the inoculum among the gonococcal isolates recovered from experimentally infected males (67–70) undergoes rapid shifts and becomes progressively more complex as infection progresses (67–70), consistent with the idea that gonococci breaching the initial barrier may encounter additional pressures (e.g., host immune responses). The phenomenon of increased phase variation as a strategy to overcome bottlenecks and regenerate population diversity has been previously described for other bacteria (71–75). Whole genome and transcriptome studies utilizing the daily gonococcal samples from the experimentally infected males of the current study to investigate phase variation and transcriptomic dynamics are an important extension of this reported study that are currently being undertaken.

In addition to the inherent limitations in characterizing the gonococcal bottleneck described above, open questions also remain with respect to the *in vivo* requirement of the MtrCDE efflux pump in human urethral infection. FA19 strains have not been characterized as a human challenge strain, and thus human experimental infection with FA19 strains were not possible to formally evaluate that the requirement of the MtrCDE pump is strain dependent in the human male urethra. It is possible that the MtrCDE pump simply does not provide an advantage in the male urethra, but confers a selective advantage in other ecologic niches, like the mouse vagina and possibly some anatomic sites in humans. By culturing urine rather than specimens directly sampled from the urethral mucosa, it is possible that the adherent infecting bacterial population is different from the bacteria that are shed and carried out of the urethra by flowing urine. Finally, whether the data of the experimental infections in males is representative of female infection or of extragenital sites remains an open question. Due to the risk of ascending gonococcal infection, experimental gonorrhea in females is not ethically acceptable. Controlled human infection models of the orpharynx and rectum do not exist yet.

Overall, our study has provided compelling information regarding the MtrCDE efflux pump, a well-studied *N. gonorrhoeae* virulence factor, and the presence of a population restriction bottleneck during *in vivo* human infection. By showing that a strain of *N. gonorrhoeae* is able to initiate a productive infection in the human male urethra even when the pump is not functional we have generated data that can inform future gonorrhea vaccine and drug development efforts that target the function of the Mtr efflux pump. By showing for the first time for a bacterial sexually-transmitetd pathogen that a bottleneck exists during *in vivo* human urethral infection we have pointed towards a gonococcal Achilles heel could be exploited for novel control strategies; knowledge of which host pressures drive the restriction of the pathogen population may answer important biological questions regarding gonococcal infection progression or reveal genes that are essential to the survival of *N. gonorrhoeae*. For example, as many as a third of all exposed individuals will not develop an infection and of the 66% of exposures that proceed to infection, ∼50% of female genital infections and ∼20% of male genital infection remain asymptomatic (data reviewed in (52)). The determinants of these phenomena are unknown and physiologic bottlenecks could play a role. Thus, the implications of our findings could have the potential of ushering in fresh lines of inquiry in our quest for new and effective anti-gonococcal strategies. This is important because in the face of increasing gonorrhea incidence and *N. gonorrhoeae’s* superbug status, the need to contain the threat of untreatable gonorrhea has become paramount.

## Methods

### Strains

*N. gonorrhoeae* strains used in this study are listed in Table 1. *N. gonorrhoeae* FA1090 served as the parent strain for the human challenge investigations, whereas wild type FA1090 and FA19 were the parent strains for the mouse challenge experiments. FA1090 is a porin serotype PIB-3, streptomycin (Sm)-resistant strain that has been used extensively in experimental human infection studies (27). FA19 has been used widely in mouse challenge studies, including studies of the Mtr efflux pump. The FA1090 human challenge inoculum strain, including the FA1090 A26 variant utilized in the current studies in men and mice is piliated and predominantly Opa-negative. FA109011*mtrD* is an isogenic mutant of FA1090, generated according to procedures described below. In addition to FA1090 A26 and FA109011*mtrD*, three other FA19 strains were used in mouse challenge experiments (FA19 wild type and two mutant strains lacking a functional Mtr pump, namely FA1911*mtrD*, and FA19 *mtrD::kan*).

### Generation and characterization of the mutant strains

To delete the *mtrD* gene and generate FA109011*mtrD,* a single colony of the FA1090 A26 as the starting material was used in a two-step transformation, as previously described (81) and used by us to delete *lptA* in FA1090 . The deletion was made without leaving behind a selectable marker (Supplementary Figure 1). Briefly, a two-gene cassette containing both a selectable marker (chloramphenicol acetyl transferase [CAT] conferring chloramphenicol [Cm] resistance) and a counter selectable marker (*rpsL*, conferring streptomycin [Sm] susceptibility on the naturally resistant FA1090) was cloned into the *mtrD* gene and used to replace the wild type gene on the chromosome by allelic exchange. A second transformation replaced the cassette-containing version of the gene with an unmarked deletion. Targeted PCR amplification of the *mtrD* locus and whole-genome sequencing of the FA109011*mtrD* genome sequence confirmed deletion of the gene in the mutant. FA1911*mtrD* was generated in the same manner as FA109011*mtrD* (i.e., leaving behind a clean deletion without a selectable marker), whereas the FA19 *mtrD::kan* mutant was constructed via an insertional mutation, leaving behind a kanamycin selectable marker, as previously described (19). While the mutant strains express the other two components of the Mtr efflux pump, the lack of *mtrD* expression leads to the inability to assemble the pump. MtrD expression contributes to gonococcal resistance to cationic antimicrobial compounds such as polymyxin B (PB). Compared to parental strain FA1090 A26 the *mtrD* deletion mutant was four-fold more PB susceptible (MICs of 100 vs. 25 µg/ml, respectively).

### Human urethral experimental infections

Procedures for human male participant recruitment, informed consent, intraurethral inoculation, an antibiotic treatment were as previously described (27, 28, 82, 83). Females are excluded from these studies due to risks of ascending gonorrhea infection into the upper reproductive tract. Between April 2017 and November 2018, separate cohorts of up to 4 participants were inoculated with FA1090 A26 alone, FA109011*mtrD* alone, or with mixtures of FA1090 A26 combined with FA109011*mtrD* containing approximately one million organisms. Gonococci in the inoculum suspensions were predominantly Opa-negative, piliated and expressed the same PilE sequence as previously characterized FA1090 variants used in experimental human infection studies(29, 68). For competitive infections in men, mixtures of wild type FA1090 A26 and FA109011*mtrD* were prepared ahead of time and stored at -≤70°C until use. Procedures pertaining to inoculum preparation for single and mixed strain infections have been previously described (27–30, 82).

Participants were observed for up to 5 days post-inoculation and received antibiotics as soon as discharge developed or on the final day of study (day 5 post-inoculation, irrespective of whether they were deemed to be infected or not). Antibiotic treatment was with cefixime (single 400 mg oral dose) or ceftriaxone (single 250 mg dose delivered intramuscularly). Infection alone without discharge did not trigger treatment. First-void urine specimens were obtained daily after inoculation for up to five days and examined quantitatively for pyuria (white blood cells) and gonococci (bacteriuria). Participants were considered to be infected if they had a positive urine culture or positive swab culture, with or without symptoms (i.e., urethral discharge). Urine sediment was cultured quantitatively on GC agar with 3 g vancomycin, 12.5 units nystatin, and 5 g trimethoprim lactate/ml, which permits the growth of FA1090 and FA109011*mtrD*. Inocula used in single and mixed strain human challenges were also plated quantitatively. Colonies were enumerated for bacterial quantitation. Up to 96 colonies per volunteer per culture day and up to 96 colonies from mixed inocula were picked and stored in freezing medium until strain determination by real-time PCR.

### Murine competitive infections

Female BALB/c mice (6 to 8 weeks old) were treated with Premarin and antibiotics to increase susceptibility to *N. gonorrhoeae* as described previously (19, 84). Mixtures of relevant strains were prepared on challenge day, as previously described (19). Groups of mice were inoculated vaginally with the following mixtures: FA1090 and FA109011*mtrD*, FA19 and FA1911*mtrD,* or FA19 and FA19 *mtrD::kan*. Vaginal swabs were collected every other day for 5 days starting on day 1 post-inoculation and suspended in 100 µl GC broth. For mixtures containing deletion mutants, vaginal swab suspensions were quantitatively cultured for *N. gonorrhoeae* on GCB plates with Sm (100 µg /ml) plus 10 µg PMB/ml. Up to 48 colonies per mouse per culture day were picked and stored in freezing medium until shipment to UNC-CH for strain determination by real-time PCR, as described below. For competitive infections with FA19 and FA19 *mtrD::kan*, vaginal swab suspensions were cultured on GC agar with Sm (100 µg /ml) for total CFU counts (wild-type and mutant) and GC agar with Sm (100 µg /ml) plus Km (50 µg/ml) to determine mutant CFU counts.

### Strain determination in human competitive infections that contained clean deletion mutants

Up to 96 colonies per human male volunteer per culture day and up to 48 colonies per mouse per culture day were individually tested for strain determination using a duplex Taqman real-time PCR assay (Supplementary Figure 1). Frozen, viable material from each colony was sub-cultured in sterile 96 well microtiter plates by adding gonococci to 75 µL of sterile GC broth (with supplements) using aseptic technique and a replica plater tool. Gonococci were expanded overnight (18-24h) at 35-37°C in a humidified atmosphere containing 5-7% CO_2_ in order to generate enough bacterial material for DNA isolation and subsequent strain detection by real time PCR.

Bacterial lysates were generated according to established procedures (85). Resulting lysates were used as DNA template in the real-time assay. In brief, each well containing bacterial material expanded from a single colony was placed in 75 µL of sterile lysis buffer containing 2 mM EDTA, 50 mM Tris Cl at pH 8.5, and 1% Triton X-100 (85). Cells were lysed in 96-well PCR plates using a thermocycler under the following cycling conditions: 15 min at 94°C followed by incubation for 5 min at 25°C. Control lysates from each individual strain were made and included as controls on each assay plate.

The simultaneous and specific detection of wild type and mutant strains was possible for three reasons (Supplementary Figure 1): 1) the wild type specific primers and probe were located within the *mtrD* gene, which is deleted in the mutant strains, and produce a 164bp product (note: the *mtrD* gene in FA1090 and FA19 have 100% genetic homology); 2) the primers and probe targeting the mutant strain were designed in a gap PCR fashion, whereby the forward and reverse primers flank the *mtrD* deletion and the probe spans either side of the deletion; only if a deletion is present, can the real-time assay utilize these primers and probe to amplify and detect the HEX-labeled 221 bp product; and 3) the two probes were labeled with two different fluorophores with non-overlapping emission spectra (wild type probe was labeled at the 5’ end with the FAM fluorophore, which has an emission spectrum of 510-530 nm; and mutant probe was labeled at the 5’ end with HEX fluorophore, which has an emission spectrum of 560-580 nm). Real-time assays were run in 384 well plates with five positive and five negative controls on each plate using a ViiA 7 Real-Time PCR System from Applied Biosystems and the following cycling conditions: 1 cycle at 95℃ for 3 min, followed by 40 cycles at each of 95℃ for 15 sec and 58℃ for 45 seconds. Primer and probe sequences, and PCR recipes are provided in Supplementary Table 3. Only wells which gave a positive amplification signal for one, but not both targets, were included for enumeration and competitive index calculation.

### Competitive index calculations

For experimental murine and human infections, results for infected individuals were expressed as a competitive index (CI) using the equation:

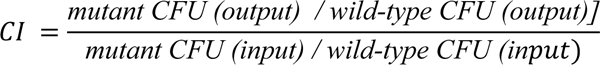

Where

Output refers to the number of wild type and mutant cfu enumerated from cultures of urine sediment or from cultures of mouse vaginal swab suspension Input refers to the number of cfu enumerated from culture of bacterial suspension used to inoculate that particular cohort/group of men or mice.

For human and mouse competitive infections, the culture limit of detection was assigned as 1 cfu/output cfu (e.g., 1/96 or 1/48, respectively). The competitive index can be equal to 1 if there is equal fitness between the two strain, greater than 1 (or greater than 0 when expressed on the logarithmic scale) if the mutant is favored or less than 1 (or less than 0 when expressed on the logarithmic scale) if the wild type is favored.

### Statistics

The evaluable population in non-competitive infections included participants who received a dose of *N. gonorrhoeae* within 1 Log_10_ of the intended dose and reached an objective study endpoint (urethral discharge or day 5). The evaluable population in competitive infections included participants who received a dose of *N. gonorrhoeae* within 1 Log_10_ of the intended dose, reached an objective study endpoint (urethral discharge or day 5) and had at least one positive urine culture.

Differences between groups were evaluated by Mann-Whitney rank-sum tests, 1-sided Fisher exact tests, or Kruskall-Wallis test, as appropriate. The CI of the mixed group was tested against the null hypothesis that the median equal zero using the one-sample Wilcoxon sign rank test and alpha of 0.05.

### Study approvals

Written informed consent from each participant was obtained prior to conducting any study activities. Experimental human infections were conducted between 2017-2019 in the Clinical and Translational Research Center of the North Carolina Translational and Clinical Sciences Institute at the University of North Carolina at Chapel Hill according to the guidelines of the U.S. Department of Health and Human Services and the University’s Institutional Review Board under a protocol for use of an investigational new drug (IND) that was authorized by the U.S. Food and Drug Administration.

All animal experiments were conducted at the Uniformed Services University of the Health Sciences according to the guidelines of the Association for the Assessment and Accreditation of Laboratory Animal Care under a protocol that was approved by the University’s Institutional Animal Care and Use Committee.

## Author contributions

MMH and JAD designed the research study, revised the manuscript, and conducted the human experimental infections in conjunction with the UNC-Global Clinical Trials Unit/DMID 09-0106 Study Team. AW performed microbiology assessments and drafted the manuscript. MMH and AW conducted data analysis. NH performed statistical analysis. WMS and JTB generated the mutant strains. AEJ, KLC, and AAB conducted all the animal challenge studies. WMS and AEJ edited the manuscript.

## Acknowledgments and Funding Information

We extend our gratitude to the study participants. James Anderson and Lorraine Balleta provided technical support for the microbiologic assessments. See Supplemental Acknowledgments for UNC-Global Clinical Trials Unit/DMID 09-0106 Study Team member details. We are grateful to Holly Baughman and Jill El-Khorazaty from Emmes for the input on the study. The authors and study were supported by funding from the National Institutes of Health, National Center for Advancing Translational Sciences, National Institute of Allergy and Infectious Diseases, and National Institute of General Medical Sciences through Grant Award Numbers U01AI114378, TL1TR002491, UL1TR001111, R01AI021150 and U19AI113170. W.M.S. is the recipient of a Senior Research Career Scientist Award from the Department of Veterans Affairs Medical Research Service.

## Data Availability

Raw data produced in the present study are available upon reasonable request to the authors or are contained in the manuscript

## Notes

Conflict-of-interest statement The authors have declared that no conflict of interest exists.

### Competing Interest Statement

The authors have declared no competing interest.

### Clinical Trial

Clinicaltrials.gov NCT03840811.

### Clinical Protocols

https://classic.clinicaltrials.gov/ct2/show/NCT03840811

### Author Declarations

Ethics committee/IRB of University of North Carolina at Chapel Hill gave ethical approval for this work under a protocol for use of an investigational new drug (IND) that was authorized by the U.S. Food and Drug Administration. The Institutional Animal Care and Use Committee of Uniformed Services University of the Health Sciences gave ethics approval for animal work according to the guidelines of the Association for the Assessment and Accreditation of Laboratory Animal Care.

## Bibliography

1. Maness MJ, and Sparling PF. Multiple antibiotic resistance due to a single mutation in Neisseria gonorrhoeae. J Infect Dis. 1973;128(3):321–30.

2. Guymon LF, Walstad DL, and Sparling PF. Cell envelope alterations in antibiotic-sensitive and-resistant strains of Neisseria gonorrhoeae. J Bacteriol. 1978;136(1):391–401.

3. Shafer WM, Qu X, Waring AJ, et al. Modulation of Neisseria gonorrhoeae susceptibility to vertebrate antibacterial peptides due to a member of the resistance/nodulation/division efflux pump family. Proc Natl Acad Sci U S A. 1998;95(4):1829–33.

4. Hagman KE, Lucas CE, Balthazar JT, et al. The MtrD protein of Neisseria gonorrhoeae is a member of the resistance/nodulation/division protein family constituting part of an efflux system. Microbiology (Reading). 1997;143 (Pt 7):2117–25.

5. Lucas CE, Hagman KE, Levin JC, et al. Importance of lipooligosaccharide structure in determining gonococcal resistance to hydrophobic antimicrobial agents resulting from the mtr efflux system. Mol Microbiol. 1995;16(5):1001–9.

6. Hagman KE, Pan W, Spratt BG, et al. Resistance of Neisseria gonorrhoeae to antimicrobial hydrophobic agents is modulated by the mtrRCDE efflux system. Microbiology (Reading). 1995;141 (Pt 3):611–22.

7. Lucas CE, Balthazar JT, Hagman KE, et al. The MtrR repressor binds the DNA sequence between the mtrR and mtrC genes of Neisseria gonorrhoeae. J Bacteriol. 1997;179(13):4123–8.

8. Warner DM, Shafer WM, and Jerse AE. Clinically relevant mutations that cause derepression of the Neisseria gonorrhoeae MtrC-MtrD-MtrE Efflux pump system confer different levels of antimicrobial resistance and in vivo fitness. Mol Microbiol. 2008;70(2):462–78.

9. Lyu M, Ayala JC, Chirakos I, et al. Structural Basis of Peptide-Based Antimicrobial Inhibition of a Resistance-Nodulation-Cell Division Multidrug Efflux Pump. Microbiol Spectr. 2022;10(5):e0299022.

10. Delahay RM, Robertson BD, Balthazar JT, et al. Involvement of the gonococcal MtrE protein in the resistance of Neisseria gonorrhoeae to toxic hydrophobic agents. Microbiology (Reading). 1997;143 (Pt 7):2127–33.

11. Handing JW, Ragland SA, Bharathan UV, et al. The MtrCDE Efflux Pump Contributes to Survival of Neisseria gonorrhoeae From Human Neutrophils and Their Antimicrobial Components. Front Microbiol. 2018;9:2688.

12. Hagman KE, and Shafer WM. Transcriptional control of the mtr efflux system of Neisseria gonorrhoeae. J Bacteriol. 1995;177(14):4162–5.

13. Shafer WM, Balthazar JT, Hagman KE, et al. Missense mutations that alter the DNA-binding domain of the MtrR protein occur frequently in rectal isolates of Neisseria gonorrhoeae that are resistant to faecal lipids. Microbiology (Reading). 1995;141 (Pt 4):907–11.

14. Janganan TK, Zhang L, Bavro VN, et al. Opening of the outer membrane protein channel in tripartite efflux pumps is induced by interaction with the membrane fusion partner. J Biol Chem. 2011;286(7):5484–93.

15. Bolla JR, Su CC, Do SV, et al. Crystal structure of the Neisseria gonorrhoeae MtrD inner membrane multidrug efflux pump. PLoS One. 2014;9(6):e97903.

16. Hoffmann KM, Williams D, Shafer WM, et al. Characterization of the multiple transferable resistance repressor, MtrR, from Neisseria gonorrhoeae. J Bacteriol. 2005;187(14):5008–12.

17. Zalucki YM, Dhulipala V, and Shafer WM. Dueling regulatory properties of a transcriptional activator (MtrA) and repressor (MtrR) that control efflux pump gene expression in Neisseria gonorrhoeae. mBio. 2012;3(6):e00446–12.

18. Rouquette C, Harmon JB, and Shafer WM. Induction of the mtrCDE-encoded efflux pump system of Neisseria gonorrhoeae requires MtrA, an AraC-like protein. Mol Microbiol. 1999;33(3):651–8.

19. Jerse AE, Sharma ND, Simms AN, et al. A gonococcal efflux pump system enhances bacterial survival in a female mouse model of genital tract infection. Infect Immun. 2003;71(10):5576–82.

20. Rouquette-Loughlin CE, Reimche JL, Balthazar JT, et al. Mechanistic Basis for Decreased Antimicrobial Susceptibility in a Clinical Isolate of Neisseria gonorrhoeae Possessing a Mosaic-Like mtr Efflux Pump Locus. mBio. 2018;9(6).

21. Veal WL, Yellen A, Balthazar JT, et al. Loss-of-function mutations in the mtr efflux system of Neisseria gonorrhoeae. Microbiology (Reading). 1998;144 (Pt 3):621–7.

22. Veal WL, Nicholas RA, and Shafer WM. Overexpression of the MtrC-MtrD-MtrE efflux pump due to an mtrR mutation is required for chromosomally mediated penicillin resistance in Neisseria gonorrhoeae. J Bacteriol. 2002;184(20):5619–24.

23. Ohneck EA, Zalucki YM, Johnson PJ, et al. A novel mechanism of high-level, broad-spectrum antibiotic resistance caused by a single base pair change in Neisseria gonorrhoeae. mBio. 2011;2(5).

24. Warner DM, Folster JP, Shafer WM, et al. Regulation of the MtrC-MtrD-MtrE efflux-pump system modulates the in vivo fitness of Neisseria gonorrhoeae. J Infect Dis. 2007;196(12):1804–12.

25. Golparian D, Shafer WM, Ohnishi M, et al. Importance of multidrug efflux pumps in the antimicrobial resistance property of clinical multidrug-resistant isolates of Neisseria gonorrhoeae. Antimicrob Agents Chemother. 2014;58(6):3556–9.

26. Ohnishi M, Golparian D, Shimuta K, et al. Is Neisseria gonorrhoeae initiating a future era of untreatable gonorrhea?: detailed characterization of the first strain with high-level resistance to ceftriaxone. Antimicrob Agents Chemother. 2011;55(7):3538–45.

27. Hobbs MM, Sparling PF, Cohen MS, et al. Experimental Gonococcal Infection in Male Volunteers: Cumulative Experience with Neisseria gonorrhoeae Strains FA1090 and MS11mkC. Front Microbiol. 2011;2:123.

28. Waltmann A DJ, Pier GB, Cywes-Bentley C, Cohen MC, Hobbs MM. In: Fabio Bagnoli GDG, Sanjay K. Phogat, Rino Rappuoli ed. Human Challenge Studies for Vaccine Development. Springer Nature; 2021.

29. Hobbs MM, Anderson JE, Balthazar JT, et al. Lipid A’s structure mediates Neisseria gonorrhoeae fitness during experimental infection of mice and men. mBio. 2013;4(6):e00892–13.

30. Jerse AE. Experimental gonococcal genital tract infection and opacity protein expression in estradiol-treated mice. Infect Immun. 1999;67(11):5699–708.

31. Feinen B, Jerse AE, Gaffen SL, et al. Critical role of Th17 responses in a murine model of Neisseria gonorrhoeae genital infection. Mucosal Immunol. 2010;3(3):312–21.

32. Jerse AE, Wu H, Packiam M, et al. Estradiol-Treated Female Mice as Surrogate Hosts for Neisseria gonorrhoeae Genital Tract Infections. Front Microbiol. 2011;2:107.

33. Jerse AE, Crow ET, Bordner AN, et al. Growth of Neisseria gonorrhoeae in the female mouse genital tract does not require the gonococcal transferrin or hemoglobin receptors and may be enhanced by commensal lactobacilli. Infect Immun. 2002;70(5):2549–58.

34. Soler-Garcia AA, and Jerse AE. Neisseria gonorrhoeae catalase is not required for experimental genital tract infection despite the induction of a localized neutrophil response. Infect Immun. 2007;75(5):2225–33.

35. Muench DF, Kuch DJ, Wu H, et al. Hydrogen peroxide-producing lactobacilli inhibit gonococci in vitro but not during experimental genital tract infection. J Infect Dis. 2009;199(9):1369–78.

36. Song W, Condron S, Mocca BT, et al. Local and humoral immune responses against primary and repeat Neisseria gonorrhoeae genital tract infections of 17beta-estradiol-treated mice. Vaccine. 2008;26(45):5741–51.

37. Leduc I, Connolly KL, Begum A, et al. The serogroup B meningococcal outer membrane vesicle-based vaccine 4CMenB induces cross-species protection against Neisseria gonorrhoeae. PLoS Pathog. 2020;16(12):e1008602.

38. Ma KC, Mortimer TD, Hicks AL, et al. Adaptation to the cervical environment is associated with increased antibiotic susceptibility in Neisseria gonorrhoeae. Nat Commun. 2020;11(1):4126.

39. Nachamkin I, Cannon JG, and Mittler RS. Monoclonal antibodies against Neisseria gonorrhoeae: production of antibodies directed against a strain-specific cell surface antigen. Infect Immun. 1981;32(2):641–8.

40. Lyu M, Moseng MA, Reimche JL, et al. Cryo-EM Structures of a Gonococcal Multidrug Efflux Pump Illuminate a Mechanism of Drug Recognition and Resistance. mBio. 2020;11(3).

41. Barnes PD, Bergman MA, Mecsas J, et al. Yersinia pseudotuberculosis disseminates directly from a replicating bacterial pool in the intestine. J Exp Med. 2006;203(6):1591–601.

42. Brown SP, Cornell SJ, Sheppard M, et al. Intracellular demography and the dynamics of Salmonella enterica infections. PLoS Biol. 2006;4(11):e349.

43. Gerlini A, Colomba L, Furi L, et al. The role of host and microbial factors in the pathogenesis of pneumococcal bacteraemia arising from a single bacterial cell bottleneck. PLoS Pathog. 2014;10(3):e1004026.

44. Grant AJ, Restif O, McKinley TJ, et al. Modelling within-host spatiotemporal dynamics of invasive bacterial disease. PLoS Biol. 2008;6(4):e74.

45. Moxon ER, and Murphy PA. Haemophilus influenzae bacteremia and meningitis resulting from survival of a single organism. Proc Natl Acad Sci U S A. 1978;75(3):1534–6.

46. Sheppard M, Webb C, Heath F, et al. Dynamics of bacterial growth and distribution within the liver during Salmonella infection. Cell Microbiol. 2003;5(9):593–600.

47. Joseph SB, Swanstrom R, Kashuba AD, et al. Bottlenecks in HIV-1 transmission: insights from the study of founder viruses. Nat Rev Microbiol. 2015;13(7):414–25.

48. Keele BF, Giorgi EE, Salazar-Gonzalez JF, et al. Identification and characterization of transmitted and early founder virus envelopes in primary HIV-1 infection. Proc Natl Acad Sci U S A. 2008;105(21):7552–7.

49. Derdeyn CA, Decker JM, Bibollet-Ruche F, et al. Envelope-constrained neutralization-sensitive HIV-1 after heterosexual transmission. Science. 2004;303(5666):2019–22.

50. Abrahams MR, Anderson JA, Giorgi EE, et al. Quantitating the multiplicity of infection with human immunodeficiency virus type 1 subtype C reveals a non-poisson distribution of transmitted variants. J Virol. 2009;83(8):3556–67.

51. Haaland RE, Hawkins PA, Salazar-Gonzalez J, et al. Inflammatory genital infections mitigate a severe genetic bottleneck in heterosexual transmission of subtype A and C HIV-1. PLoS Pathog. 2009;5(1):e1000274.

52. Lovett A, and Duncan JA. Human Immune Responses and the Natural History of Neisseria gonorrhoeae Infection. Front Immunol. 2018;9:3187.

53. Ercoli G, Fernandes VE, Chung WY, et al. Intracellular replication of Streptococcus pneumoniae inside splenic macrophages serves as a reservoir for septicaemia. Nat Microbiol. 2018;3(5):600–10.

54. Kono M, Zafar MA, Zuniga M, et al. Single Cell Bottlenecks in the Pathogenesis of Streptococcus pneumoniae. PLoS Pathog. 2016;12(10):e1005887.

55. Zafar MA, Kono M, Wang Y, et al. Infant Mouse Model for the Study of Shedding and Transmission during Streptococcus pneumoniae Monoinfection. Infect Immun. 2016;84(9):2714–22.

56. Liu X, Kimmey JM, Matarazzo L, et al. Exploration of Bacterial Bottlenecks and Streptococcus pneumoniae Pathogenesis by CRISPRi-Seq. Cell Host Microbe. 2021;29(1):107–20 e6.

57. Hadad R, Jacobsson S, Pizza M, et al. Novel meningococcal 4CMenB vaccine antigens - prevalence and polymorphisms of the encoding genes in Neisseria gonorrhoeae. APMIS. 2012;120(9):750–60.

58. Marjuki H, Topaz N, Joseph SJ, et al. Genetic Similarity of Gonococcal Homologs to Meningococcal Outer Membrane Proteins of Serogroup B Vaccine. mBio. 2019;10(5).

59. Perrin A, Bonacorsi S, Carbonnelle E, et al. Comparative genomics identifies the genetic islands that distinguish Neisseria meningitidis, the agent of cerebrospinal meningitis, from other Neisseria species. Infect Immun. 2002;70(12):7063–72.

60. Tinsley CR, and Nassif X. Analysis of the genetic differences between Neisseria meningitidis and Neisseria gonorrhoeae: two closely related bacteria expressing two different pathogenicities. Proc Natl Acad Sci U S A. 1996;93(20):11109–14.

61. Bayliss CD, Hoe JC, Makepeace K, et al. Neisseria meningitidis escape from the bactericidal activity of a monoclonal antibody is mediated by phase variation of lgtG and enhanced by a mutator phenotype. Infect Immun. 2008;76(11):5038–48.

62. Banerjee A, Wang R, Uljon SN, et al. Identification of the gene (lgtG) encoding the lipooligosaccharide beta chain synthesizing glucosyl transferase from Neisseria gonorrhoeae. Proc Natl Acad Sci U S A. 1998;95(18):10872–7.

63. Shafer WM, Datta A, Kolli VS, et al. Phase variable changes in genes lgtA and lgtC within the lgtABCDE operon of Neisseria gonorrhoeae can modulate gonococcal susceptibility to normal human serum. J Endotoxin Res. 2002;8(1):47–58.

64. Gulati S, Zheng B, Reed GW, et al. Immunization against a saccharide epitope accelerates clearance of experimental gonococcal infection. PLoS Pathog. 2013;9(8):e1003559.

65. Gulati S, Beurskens FJ, de Kreuk BJ, et al. Complement alone drives efficacy of a chimeric antigonococcal monoclonal antibody. PLoS Biol. 2019;17(6):e3000323.

66. Golubchik T, Batty EM, Miller RR, et al. Within-host evolution of Staphylococcus aureus during asymptomatic carriage. PLoS One. 2013;8(5):e61319.

67. Jerse AE, Cohen MS, Drown PM, et al. Multiple gonococcal opacity proteins are expressed during experimental urethral infection in the male. J Exp Med. 1994;179(3):911–20.

68. Seifert HS, Wright CJ, Jerse AE, et al. Multiple gonococcal pilin antigenic variants are produced during experimental human infections. J Clin Invest. 1994;93(6):2744–9.

69. Wright CJ, Jerse AE, Cohen MS, et al. Nonrepresentative PCR amplification of variable gene sequences in clinical specimens containing dilute, complex mixtures of microorganisms. J Clin Microbiol. 1994;32(2):464–8.

70. Hamrick TS, Dempsey JAF, Cohen MS, et al. Antigenic variation of gonococcal pilin expression in vivo: analysis of the strain FA1090 pilin repertoire and identification of the pilS gene copies recombining with pilE during experimental human infection. Microbiology (Reading). 2001;147(Pt 4):839–49.

71. Gor V, Ohniwa RL, and Morikawa K. No Change, No Life? What We Know about Phase Variation in Staphylococcus aureus. Microorganisms. 2021;9(2).

72. Aidley J, Rajopadhye S, Akinyemi NM, et al. Nonselective Bottlenecks Control the Divergence and Diversification of Phase-Variable Bacterial Populations. mBio. 2017;8(2).

73. Libby E, and Rainey PB. Exclusion rules, bottlenecks and the evolution of stochastic phenotype switching. Proc Biol Sci. 2011;278(1724):3574–83.

74. Palmer ME, Lipsitch M, Moxon ER, et al. Broad conditions favor the evolution of phase-variable loci. mBio. 2013;4(1):e00430–12.

75. Moxon R, and Kussell E. The impact of bottlenecks on microbial survival, adaptation, and phenotypic switching in host-pathogen interactions. Evolution. 2017;71(12):2803–16.

76. Abraham SN, and Miao Y. The nature of immune responses to urinary tract infections. Nat Rev Immunol. 2015;15(10):655–63.

77. Pudney J, and Anderson D. Innate and acquired immunity in the human penile urethra. J Reprod Immunol. 2011;88(2):219–27.

78. Abel S, Abel zur Wiesch P, Davis BM, et al. Analysis of Bottlenecks in Experimental Models of Infection. PLoS Pathog. 2015;11(6):e1004823.

79. Qu J, Prasad NK, Yu MA, et al. Modulating Pathogenesis with Mobile-CRISPRi. J Bacteriol. 2019;201(22).

80. Garvin LE, Bash MC, Keys C, et al. Phenotypic and genotypic analyses of Neisseria gonorrhoeae isolates that express frequently recovered PorB PIA variable region types suggest that certain P1a porin sequences confer a selective advantage for urogenital tract infection. Infect Immun. 2008;76(8):3700–9.

81. Johnston DM, and Cannon JG. Construction of mutant strains of Neisseria gonorrhoeae lacking new antibiotic resistance markers using a two gene cassette with positive and negative selection. Gene. 1999;236(1):179–84.

82. Cohen MS, Cannon JG, Jerse AE, et al. Human experimentation with Neisseria gonorrhoeae: rationale, methods, and implications for the biology of infection and vaccine development. J Infect Dis. 1994;169(3):532–7.

83. Cornelissen CN, Kelley M, Hobbs MM, et al. The transferrin receptor expressed by gonococcal strain FA1090 is required for the experimental infection of human male volunteers. Mol Microbiol. 1998;27(3):611–6.

84. Raterman EL, and Jerse AE. Female Mouse Model of Neisseria gonorrhoeae Infection. Methods Mol Biol. 2019;1997:413–29.

85. Dillard JP. Genetic Manipulation of Neisseria gonorrhoeae. Curr Protoc Microbiol. 2011;Chapter 4:Unit4A 2.

86. Mickelsen PA, and Sparling PF. Ability of Neisseria gonorrhoeae, Neisseria meningitidis, and commensal Neisseria species to obtain iron from transferrin and iron compounds. Infect Immun. 1981;33(2):555–64.

