## Supplement for "*Neisseria gonorrhoeae* MtrCDE Efflux Pump During *In Vivo* Experimental Genital Tract Infection in Men and Mice Reveals the Presence of Within-Host Colonization Bottleneck"

[1] Division of Infectious Diseases, Department of Medicine, School of Medicine, University of North Carolina, Chapel Hill, NC, United States
[2] Department of Microbiology and Immunology, Emory University School of Medicine, Atlanta, Georgia, United States
[3] Department of Microbiology and Immunology, Uniformed Services University, Bethesda, Maryland, United States
[4] The Emmes Corporation, Rockville, Maryland, United States
[5] The Emory Antibiotic Resistance Center, Emory University School of Medicine, Atlanta, Georgia, United States
[6] Laboratories of Bacterial Pathogenesis, Veterans Affairs Medical Center (Atlanta), Decatur, Georgia, United States
[7] See Supplemental Acknowledgments for , UNC-Global Clinical Trials Unit/DMID 09-0106 Study Team member details
[8] Department of Microbiology and Immunology, University of North Carolina at Chapel Hill, Chapel Hill, North Carolina, United States
[9] Lineberger Comprehensive Cancer Center, University of North Carolina at Chapel Hill, Chapel Hill, NC, United States
[10] Department of Pharmacology, School of Medicine, University of North Carolina, Chapel Hill, NC, United States
* Co-senior authors contributed equally
Address correspondence to: Joseph A. Duncan, Division of Infectious Diseases (Campus Box 7030), 130 Mason Farm Road, Chapel Hill, NC, 27599-7030, USA.; Phone: 1 (919) 843-0715

Supplemental Acknowledgments

UNC-Global Clinical Trials Unit/DMID 09-0106 Study Team member details:
Susan Blevins, ACNP – Clinical Research Coordinator
Brian Gurney, ACNP – Clinical Research Coordinator
Dania Munson – Regulatory Coordinator
Brittney Soderman – Clinical Research Assistant
Catherine Kronk – Data Manager

A.

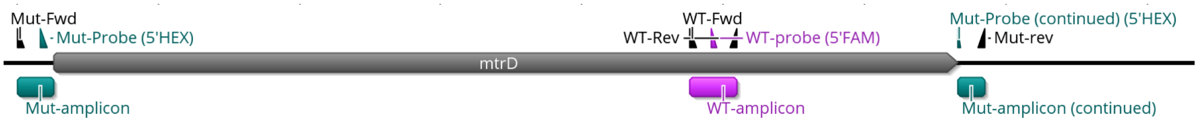


B.

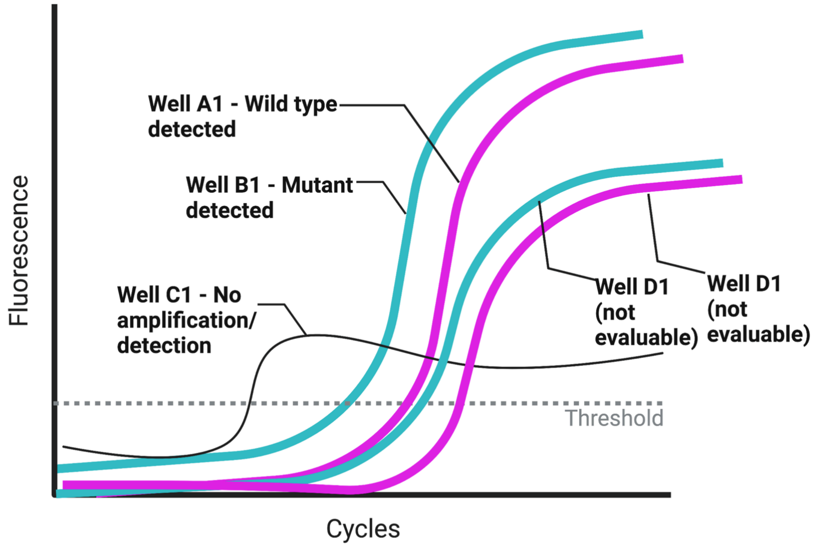


**<u>Supplementary Figure 1.</u>** Schematic of the duplex Taqman real-time PCR assay design used to distinguish between wild type and mutant FA1090 and FA19 strains (A) and theoretical detection scenarios (B). In both panels, the wild type specific probe (5'FAM labeled), amplicon, and FAM-based detection are shown in pink. The mutant specific probe (5'HEX), amplicon and HEX-based detection are shown in teal. The *mtrD* gene in FA1090 and FA19 have 100% genetic homology)

A). The wild type specific primers and probe were purposefully designed to be located within the *mtrD* gene, which is deleted in the mutant strains; the resulting amplicon is a 164bp product that emits FAM fluorescence. The primers and probe targeting the mutant strain were designed in a gap PCR fashion, whereby the forward and reverse primers flank the *mtrD* deletion and the probe spans either side of the deletion; only if a deletion is present, can the real-time assay make use of these primers and probe to amplify and detect the 221 bp product, which emits HEX fluorescence with each round of amplification. In the absence of the gene deletion (i.e., if a wild type strain were present), the resulting product would be approximately 3,000 base pirs long, whose amplification efficiency with real-time chemistry is virtually nil, and thus no HEX-based fluorescence is detected.

B). Only wells with detection signal for either FAM (wild type specific amplification) or HEX (mutant specific amplification), but not both, were evaluable and were included in strain determination analyses. Wells in which both fluorophores were detected or had no detectable/amplified PCR product were deemed not evaluable and were not included in strain determination analyses.

**Supplementary Table 1.** Real-time PCR primers, probes, and reaction recipes

| **Component** | **Sequence** | **Working concentration (µM)** | **Volume per reaction (µL)** | **Final concentration per reaction (µM)** |
|---|---|---|---|---|
| Bio-Rad iQ Multiplex Powermix | N/A | 2x | 6 | 1x |
| mtrD-WT-Taq-Fwd | TCGTCTTATGTCAGCGACTTC | 10µM | 0.6 | 0.5 |
| mtrD-WT-Taq-Rev | TCCCAAGAAACAGTAGCAATG | 10µM | 0.6 | 0.5 |
| mtrD-mut-Taq-Fwd | CGAAAAAGGTAACGCCTAAAG | 10µM | 0.6 | 0.5 |
| mtrD-mut-Taq-Rev | CTGCAACAGAGGTCAAGGTAG | 10µM | 0.6 | 0.5 |
| mtrD-WT-Taq-probe | 5'FAM-GTATGCAGCCTGCCGATATT-3'BHQ-1 | 10µM | 0.25 | 0.2 |
| mtrD-mut-Taq-probe | 5'HEX-AAGCCAAACCTGCTTCTGAA-3'BHQ-1 | 10µM | 0.25 | 0.2 |
| Water | N/A | - | 1.1 | - |

**Supplementary Table 2.** Strain composition of the gonococcal population in mixed inocula and of gonococci recovered from first void urine of men from treatment day. The participants shown in grey were not evaluable and their data was not included in final analyses.

| Cohort | Participant ID[a] | % FA1090Δ*mtrD* in inoculum | % FA1090 in inoculum | Infected? | % (n[b]) FA1090Δ*mtrD* recovered on Tx day | % (n[b]) FA1090 recovered on Tx day | Included in final analyses |
|---|---|---|---|---|---|---|---|
| 1 | *280* | *58.0* | *42.0* | *Not evaluable* | *NA* | *NA* | *No* |
| | 281 | | | Y | 0 (0) | 100 (96) | Yes |
| | *282* | | | *Not evaluable* | *NA* | *NA* | *No* |
| 2 | 291 | 54.9 | 45.1 | Y | 100 (96) | 0 (0) | Yes |
| | 292 | | | Y | 100 (75) | 0 (0) | Yes |
| 3 | 300 | 55.4 | 44.6 | Y | 0 | 100 (93) | Yes |
| | 301 | | | Y | 0 | 100 (87) | Yes |
| | 302 | | | Y | 8.9 (7) | 91.1 (72) | Yes |
| | 303 | | | Y | 100 (90) | 0 (0) | Yes |
| 4 | 311 | 75.5 | 24.5 | Y | 0 (0) | 100 (40) | Yes |
| | 313 | | | Y | 100 (96) | 0 (0) | Yes |
| | 314 | | | Y | 100 (93) | 0 (0) | Yes |

[a]All participants. Urine culture negative participants at day 5 after inoculation are shown in grey italics.

[b]Number of evaluable cfu

[c]This participant was urine culture-negative throughout the study

[d]This participant did not comply with the first void urine requirement for bacterial cultures

**Supplementary Table 3.** Strain composition of the gonococcal population in mixed inocula containing wild type FA1090 and FA1090Δ*mtrD* that was given to mice and of gonococci recovered from mouse genital swab cultures from the final positive culture day.

| Group | Mouse | % mutant CFU recovered from inoculum | % wild-type CFU recovered from inoculum | n (%) mutant recovered on final day | n (%) wild-type recovered on Tx day |
|---|---|---|---|---|---|
| 1 | 1 | 60.0 | 40.0 | 0 (0) | 90 (100) |
| | 2 | | | 0 (0) | 91 (100) |
| | 3 | | | 69 (80.2) | 17 (19.8) |
| | 4 | | | 77 (81.9) | 17 (18.1) |
| 2 | 5 | 39.6 | 60.4 | 35 (100) | 0 (0) |
| | 6 | | | 41 (97.6) | 1 (2.4) |
| | 7 | | | 0 (0) | 40 (100) |
| | 8 | | | 41 (93.2) | 3 (6.8) |
| | 9 | | | 12 (30.8) | 27 (69.2) |
| | 10 | | | 3 (6.7) | 42 (93.3) |
| | 11 | | | 45 (100) | 0 (0) |
